# Effectiveness of Non-pharmaceutical Interventions to Contain COVID-19: A Case Study of the 2020 Spring Pandemic Wave in New York City

**DOI:** 10.1101/2020.09.08.20190710

**Authors:** Wan Yang, Jaimie Shaff, Jeffrey Shaman

## Abstract

As COVID-19 continues to pose significant public health threats, quantifying the effectiveness of different public health interventions is crucial to inform intervention strategies. Using detailed epidemiological and mobility data available for New York City and comprehensive modeling accounting for under-detection, we reconstruct the COVID-19 transmission dynamics therein during the 2020 spring pandemic wave and estimate the effectiveness of two major non-pharmaceutical interventions—lockdown-like measures that reducing contact rates and universal masking. Lockdown-like measures were associated with >50% transmission reduction for all age groups. Universal masking was associated with a ∼7% transmission reduction overall and up to 20% reduction for 65+ year-olds during the first month of implementation. This result suggests that face covering can substantially reduce transmission when lockdown-like measures are lifted but by itself may be insufficient to control SARS-CoV-2 transmission. Overall, findings support the need to implement multiple interventions simultaneously to effectively mitigate COVID-19 spread before the majority of population can be protected through mass-vaccination.

## Introduction

Since the emergence of SARS-CoV-2 in late 2019, the virus has infected over 79 million people and killed over 1.75 million worldwide by the end of 2020 (as of 12/27/20).^1^ Non-pharmaceutical interventions such as social distancing and face covering have been the main strategies to contain COVID-19 during this pandemic in 2020. In late 2020, Phase-III trials for several SARS-CoV-2 vaccines showed highly promising results and were granted emergency use in several countries.^2,3^ However, before these vaccines become widely available to the general population (likely in mid-or late 2021), non-pharmaceutical interventions will need to remain the main strategies to contain COVID-19. In addition, future (re)emerging infectious disease outbreaks may need to rely on similar non-pharmaceutical measures. It is thus critical to understand the effectiveness of different non-pharmaceutical interventions implemented during the COVID-19 pandemic waves in order to inform effective future planning while balancing economic need. For instance, with face covering and social distancing by closing businesses as two main interventions, the more effective face covering is, the more businesses could remain open. As a simplified calculation, with a basic reproductive number (*R*_*0*_) of 3 and minimal immunity, a city could maintain 55% business capacity while curbing epidemic growth, if its residents could reduce transmission by 40% using face covering [ie, effective reproductive number *R*_*t*_ = 3 × 55% × (1 – 40%) = 0.99 < 1]; this threshold business capacity would drop to 33% if no residents used face covering.

However, assessing the effectiveness and impact of a given intervention for COVID-19 has been challenging due to low infection detection rates (many asymptomatic and mild infections do not seek care or receive testing),^4^ fluctuation of those infection detection rates, differential disease manifestation by age group,^5,6^ and concurrent public health interventions. As such, while a few studies have assessed the overall effectiveness of lockdown-like measures, to date, the effectiveness of specific measures including face covering *under real-world conditions* remains unclear.

To estimate the effectiveness of different non-pharmaceutical public health interventions, here we thus focus on the 2020 spring COVID-19 pandemic wave in New York City (NYC), the first COVID-19 epidemic center in the United States, where detailed data are also available. NYC experienced widespread COVID-19 transmission citywide since early March and recorded over 200,000 cases and over 21,000 COVID-19 confirmed or probable deaths during the following three months. To curb this intense transmission, NY State and NYC implemented multiple intervention measures, including health promotion campaigns in early March, telecommuting and staggered work schedule recommendations beginning the week of March 8, public schools closure starting the week of March 15,^7^ stay-at-home orders for non-essential workers starting the week of March 22,^8^ and requirements for use of face covering in public starting the week of April 12.^9^ With these overlapping and far reaching public health interventions, case diagnoses and hospitalizations peaked in April, and started to decline substantially in late April and May. NYC was able to begin its phased re-opening of industries starting the week of June 7, 2020.

In this study, we apply a model-inference system^10-12^ developed to support the city’s COVID-19 pandemic response to reconstruct the underlying transmission dynamics of COVID-19 in NYC during March 1 – June 6, 2020 (i.e. prior to the city’s reopening). To address the aforementioned challenges, our model-inference system simultaneously assimilates three sources of data: 1) confirmed COVID-19 case data, 2) COVID-19 associated death data (both cases and deaths are assimilated by neighborhood and age group), and 3) neighborhood-level mobility data to constrain the model system. This enables inference of the overall infection rate (i.e. including those not documented by surveillance), estimation of key transmission characteristics (e.g., the reproductive number) through time, and assessment of the effectiveness of different public health interventions, including social distancing and face covering, implemented over time. We further incorporate these estimates to project cases and deaths in the weeks beyond our study period and compare the projections to independent observations in order to evaluate the accuracy of these estimates. We conclude with a discussion on the implication of our findings on strategies to safely reopen economies in places COVID-19 continues to pose substantial public health threats.

## Results

### Overall Epidemic Trends

Following diagnosis of the first case in NYC, confirmed COVID-19 cases in the entire population increased nearly exponentially during the first three weeks (Fig 1B) before slowing down beginning the week of March 22, 2020 when NYC implemented a stay-at-home order. However, case trajectories differed substantially by age group. Foremost, reported cases increased with age: the case trajectory for those aged 25-44 years mirrored the overall epidemic curve, those older than 45 years had higher case rates, and those under 25 years had the lowest case rates (Fig 1A and Table S1). Infants (i.e. <1 year), however, had higher case rates than 1-4 and 5-14 year-olds (Fig 1). In addition, the timing of peak case rate was mixed. Case rates in 25-44, 45-64, and 65-74 year-olds peaked earliest during the week of March 29, 2020, followed by <1, 1-4, 15-24, and 75+ year-olds with a 1-week lag; in comparison, the case rate for 5-14 year-olds fluctuated with a less clear peak during the weeks of March 29 – April 26, 2020.

**Fig 1.**
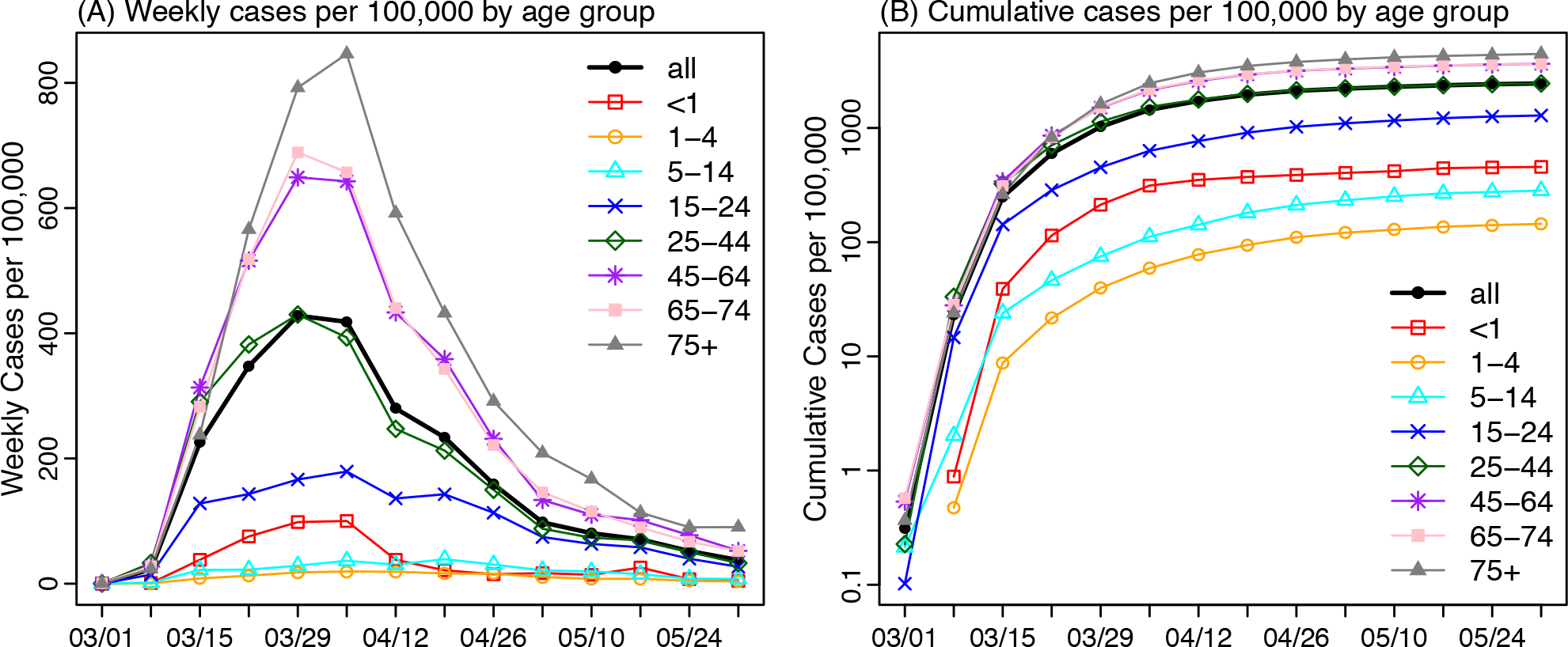
Epidemic dynamics. Reported laboratory confirmed cases (A) and cumulative cases (B) per 100,000 population by week of diagnosis for all ages overall and by age group.

The epidemic trends based on diagnosed cases, however, were obscured by varying infection detection rates by age and through time. COVID-19 infections are more likely to manifest as symptomatic illness and/or more severe disease in individuals with underlying conditions and in older adults.^5,6^ Such differential clinical characteristics by age thus lead to varying healthcare seeking behaviors and infection detection rates by age. In addition, testing policies varied over the course of the Spring 2020 COVID-19 pandemic in NYC. During this time, testing capacity was limited at the federal, state and local levels by guidelines for who should be tested (due, for example, to limited availability of test kits, swabbing supplies and reagents), which required prioritizing testing for severely ill patients and those highly vulnerable to severe disease. Testing capacities expanded during the week of March 8, 2020;^13^ however, by late March, material shortages (including testing kits and personal protective equipment) again prompted the city to restrict testing to those severely ill.^14^ Using our model-inference system, we estimated that infection detection rates increased in early March, reaching a peak of around 20% for all ages overall during the week of March 15, and declined afterwards before increasing again in mid-April.^11,12^

After accounting for infection detection rates to include undiagnosed infections, a different picture of the NYC spring outbreak emerges (Fig 2 and Fig S3). Estimated infection rates were highest among 25-44 and 45-64 year-olds (Fig. 2 F and G), followed by 65-74 and 75+ year-olds (Fig. 2 H and I), then 5-14 and 15-24 year-olds (Fig. 2 D and E), and were lowest among <1 and 1-4 year olds (Fig. 2 B and C; Table S1). Estimated infection rates in the younger age groups (in particular, 5-14, 15-24, and 25-44 year-olds; Fig. 2 D-F) peaked during the week of March 22, 2020, followed by the three older age groups (i.e. 45-64, 65-74, and 75+ year-olds; Fig. 2 G-I) about a week later.

**Fig 2.**
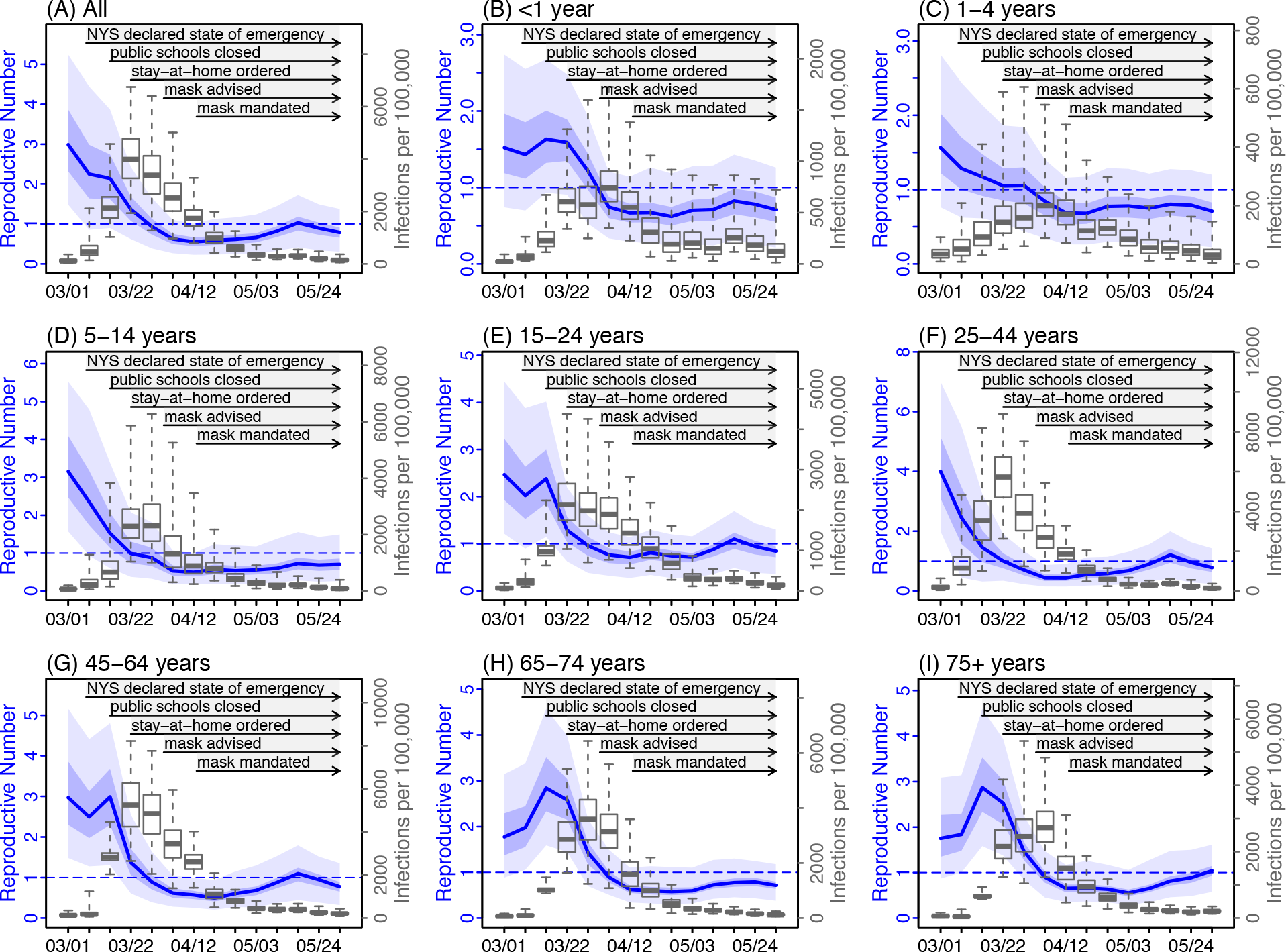
Estimated changes in the effective reproductive number and infection rates. Blue lines show the estimated effective reproductive number (*R*_*t*_) for each week; surrounding areas show the 50% and 95% CrIs. Superimposed boxes (right y-axis) show estimated infection rates by week: median (thick vertical lines), 50% CrIs (box edges), and 95% CrIs (whiskers).

Given the large uncertainties in model estimates, we verified our estimates of infection rates using available serology data collected during three phases of the pandemic (i.e. early-phase in March,^15^ mid-phase in April,^16^ and end of the pandemic wave in June^17^). Overall, our estimated cumulative infection rates were in line with corresponding measures from antibody tests, for all three phases of the pandemic wave (for details, see the Appendix of Yang et al. 2020^11,12^)

### Overall effectiveness of interventions

The reproductive number at time-*t* (*R*_*t*_) measures the average number of persons an infected individual infects and thus reflects underlying epidemic dynamics. The epidemic expands in size if *R*_*t*_ is above unity and subsides otherwise. In addition, when the entire population is susceptible and no interventions are in place, *R*_*t*_, referred to as the basic reproductive number (*R*_*0*_), reflects the transmissibility of an infection in that population. Here we estimated that *R*_*t*_ was 2.99 [median and interquartile range (IQR): 2.32 – 3.86; Table S2] during the first week of the pandemic (i.e. the week of March 1) in NYC, similar to *R*_*0*_ estimates reported for other places.^18,19^ It decreased to around 2.2 during the next two weeks, when NY State declared a state of emergency and public awareness and voluntary precautionary measures (e.g. avoiding public transit^20^) increased (Fig 2A). Following the stay-at-home mandate starting the week of March 22, *R*_*t*_ dropped substantially to 1.37 (IQR: 1.08 – 1.68) during that first week, to 0.93 (IQR: 0.73 – 1.13) a week later, and to a minimum of 0.56 (IQR: 0.45 – 0.67) during the week of April 12 (Fig 2A). These prompt decreases in *R*_*t*_ from mid-March to mid-April indicate that implemented public health messaging and interventions were effective in curtailing COVID-19 transmission.

Similar decreases in *R*_*t*_ occurred among most age groups (Fig 2 B-I). Overall, *R*_*t*_ among younger age groups (<45 years) decreased one or two weeks earlier than older age groups (45-64, 65-74, and 75+; Fig 2 C-F vs. Fig 2 G-I). Of note, among the four age groups with higher contact rates^21^ (i.e. 5-14, 15-24, 25-44, and 45-64 year-olds), *R*_*t*_ dropped below 1 the earliest among 5-14 year-olds (0.99, IQR: 0.74 – 1.30; Table S2) during the week of March 22. This is consistent with the earliest public health interventions to this age group: the closure of public schools beginning the week of March 15.^7^

### Effectiveness of reducing contact via school closure and voluntary or mandated stay-at-home measures

Several public health interventions were implemented around the same time (Fig. 2), and some interventions may take longer to produce an effect (e.g., due to slower compliance with the measure). It is thus challenging to separate the impact of different interventions. However, a number of measures – including voluntarily working from home during the early weeks of the pandemic, school closures, and the stay-at-home mandate – in effect reduce rates of close in-person contact, a key factor for COVID-19 transmission. Thus, here we focus on estimating the impact of interventions whose primary mechanism of action is through a reduction in population contact rates. Given the difficulties measuring this quantity directly, we instead approximated population contact rates using human mobility data, which record real time population movement based on location changes of individual mobile devices (see Data). Indeed, the reduction in *R*_*t*_ mirrored the reduction in mobility (Fig 3). The Pearson correlation (*r*) between *R*_*t*_ and mobility over the 14-week study period was 0.96 for all ages overall and ≥0.9 for 1-4, 5-14, 15-24, and 25-44 year-olds (Table S3). Thus, we focused on mobility as a proxy for contact rates and used this quantity to estimate the corresponding changes in *R*_*t*_ and segregate the impact of interventions that reduce population contact rates from other concurrent interventions.

**Fig 3.**
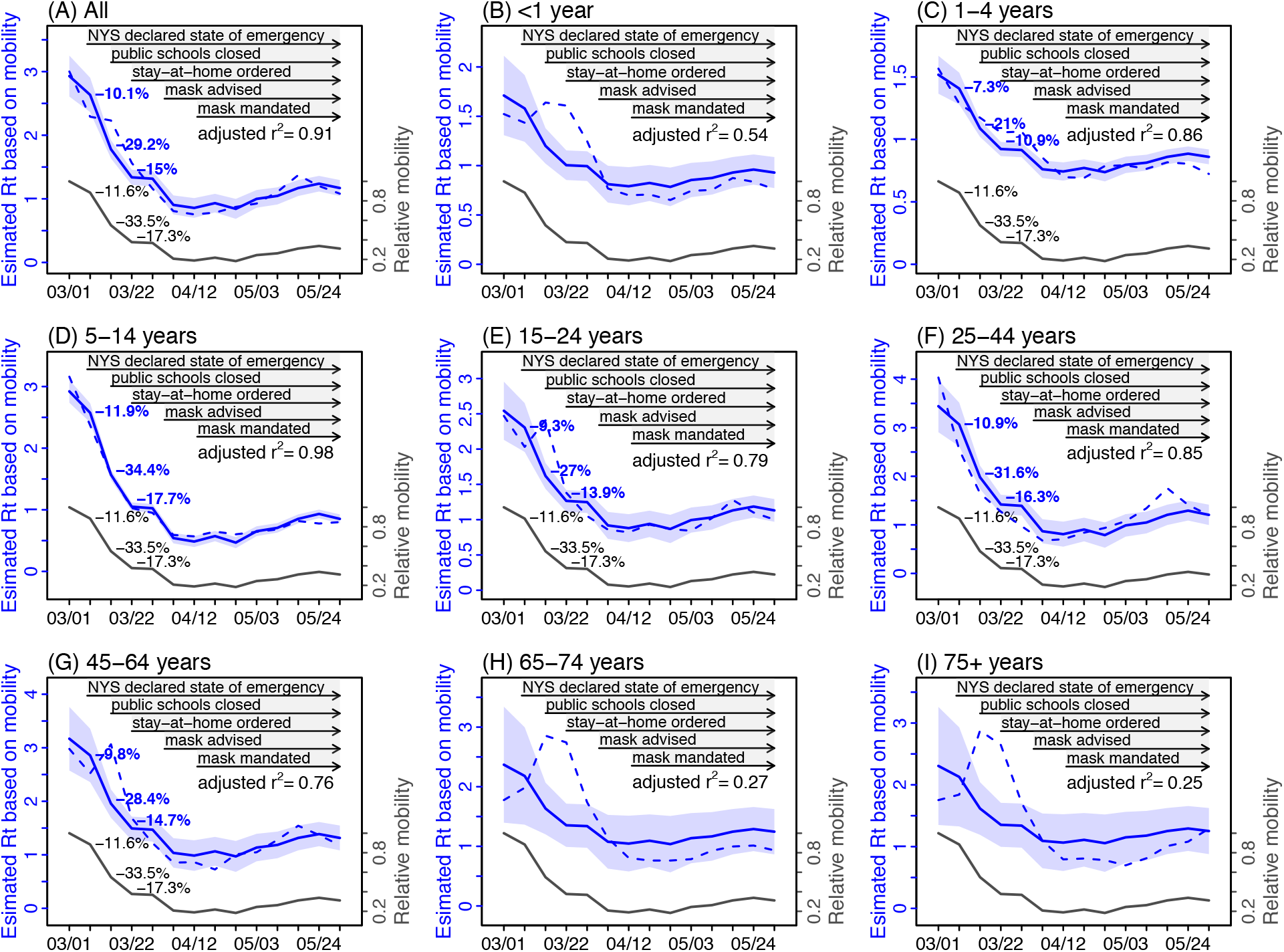
Effectiveness of reducing contact rates. Note the estimated effectiveness combined all measures that reduce contact rates, including school closures and voluntary or mandated stay-at-home measures. Dark grey lines show the observed changes in mobility (right y-axis). Blue lines show *R*_*t*_ estimated using a linear regression model with mobility as the sole predictor; surrounding areas show the 95% CrIs of the model estimates. The adjusted r^2^ for the regression model is also shown in each plot. For comparison, dashed blue lines show *R*_*t*_ estimates from the model-inference system, without accounting for susceptibility. Percentages attached to the lines show the incremental reductions in either estimated *R*_*t*_ (in blue) or mobility (in grey).

Mobility reduced by 11.6% during the second week of the pandemic in NYC (i.e. the week of March 8), and by a further 33.5% and 17.3% in the following two weeks, respectively (Fig 3). Using observed mobility data (i.e. our proxy for population contact rates) to estimate the corresponding changes in *R*_*t*_, we estimate that, for all ages overall, reductions in population contact rates were associated with *R*_*t*_ reductions of 10.1% (95% CI: 8.3 – 11.9%) by the second week of the pandemic, and another 29.2% (95% CI: 24.9 – 33.5%) and 15.0% (95% CI: 14.3 – 15.8%) in the following two weeks, respectively (Fig 3A). By the week of April 12 when *R*_*t*_ reached its minimum, the reduction in population contact rates was associated with an *R*_*t*_ reduction of 70.7% (95% CI: 65.0 – 76.4%). In addition, analysis at the neighborhood level consistently showed large reductions in *R*_*t*_ that were likely due to reductions in population contact rates (range of median estimates: 66.1 – 90.1% across the 42 neighborhoods in NYC; Fig S4).

In addition, transmission in four age groups (i.e., 5-14, 15-24, 25-44, and 45-64 year-olds) appeared to be most impacted by changing population contact rates (Fig 3 and Table 1). The reduction in population contact rates was associated with decreases of the age-specific *R*_*t*_ by 83.4% (95% CI: 80.1 – 86.7%) for 5-14 year-olds, 65.4% (95% CI: 57.0 – 73.8%) for 15-24 year-olds, 76.5% (95% CI: 68.5 – 84.6%) for 25-44 year-olds, and 68.9% (95% CI: 59.2 – 78.6%) for 45-64 year-olds, by the week of April 12.

**Table 1.** Estimated effectiveness of reducing contact rate and face covering. Numbers are the estimated mean and 95% CIs, in percentage. Note the estimated effectiveness of contact rate reduction combined all measures that reduce contact rates, including school closures and voluntary or mandated stay-at-home measures.

|  | Estimated effectiveness of intervention (%) |  |  |
| --- | --- | --- | --- |
|  | contact rate reduction | Face covering (1<sup>st</sup> month) | Face covering (2 months) |
| Age |  |  |  |
| all | 70.7 (65.0, 76.4) | 6.6 (0.8, 12.4) | 3.4 (-1.9, 8.6) |
| <1 | 53.8 (41.6, 66) | 9.3 (-4.2, 22.9) | 12.8 (0.2, 25.3) |
| 1-4 | 51.0 (45.8, 56.2) | 0.9 (-5.5, 7.4) | 6.7 (0.6, 12.8) |
| 5-14 | 83.4 (80.1, 86.7) | 3.0 (-0.5, 6.6) | 1.6 (-1.6, 4.8) |
| 15-24 | 65.4 (57.0, 73.8) | 4.3 (-2.8, 11.4) | 4.0 (-2.5, 10.6) |
| 25-44 | 76.5 (68.5, 84.6) | 4.5 (-0.6, 9.7) | -1.0 (-5.6, 3.7) |
| 45-64 | 68.9 (59.2, 78.6) | 8.1 (-0.1, 16.1) | 4.4 (-2.9, 11.8) |
| 65-74 | 55.8 (34.5, 77.2) | 20.8 (-0.1, 41.6) | 18.3 (-0.2, 36.9) |
| 75+ | 53.8 (32.3, 75.3) | 20.8 (-0.9, 42.5) | 16.2 (-3.3, 35.7) |

### Effectiveness of face covering/masking under real-world conditions

Estimated transmission rates (or probability of infection) and the infectious period also closely tracked changes in mobility (Table S3; *r*≥0.5 for most age groups). Thus, it appears that reducing mobility not only reduces contact rates but also likely reduces 1) the probability of transmission per contact due to, e.g., increased public spacing and 2) the effective infectious period per infected individual due to, e.g., more time spent at home and as a result reduced time for community transmission despite likely unchanged duration of viral shedding. Given this observation, we hypothesize that the relationship between mobility and estimated transmission rates (and effective infectious period) can be used to disentangle the impact of interventions that reduce population contact rates (particularly, the stay-at-home mandate) and face covering/masking — two major public health interventions implemented in NYC during the pandemic — on transmission. We make two predictions if this hypothesis holds. First, predicted transmission rates (infectious period) using mobility data alone would be higher (longer) than those estimated by the model-inference system additionally based on case and mortality data *for weeks when face covering in public was mandated* as it would lead to further reductions in transmission (i.e., temporality and direction of the impact). Second, while the efficacy of masking (i.e., measured under ideal conditions of mask quality and correct use) likely does not vary by individual, the effectiveness of masking (i.e., measured under real-world, often imperfect conditions) and impact of this intervention could vary by subpopulation due, for example, to different usage rates of masks; as such, we expect the predictive errors to be larger for age groups with higher compliance of masking (i.e., magnitude of the impact). Our analyses largely confirmed both predictions. For the first, as shown in Fig 4, transmission rates predicted using a linear regression model with the observed mobility as the sole predictor were higher than those estimated by the model-inference system, following the face covering mandate starting the week of April 12.^9^ For the latter, the discrepancies in the two model estimates (i.e. the gaps between the dashed and solid blue lines; Fig 4) appeared to increase with age and were largest among the two elderly age groups who have been reported to more frequently use masks.^22-24^ However, infants (<1 year) appeared to have a larger reduction than other children groups; this could have been due to transmission reduction related to their sources of infection (e.g. their caretakers and healthcare settings where they tended to be exposed). Similar patterns held for the effective infectious period (Fig 4).

**Fig 4.**
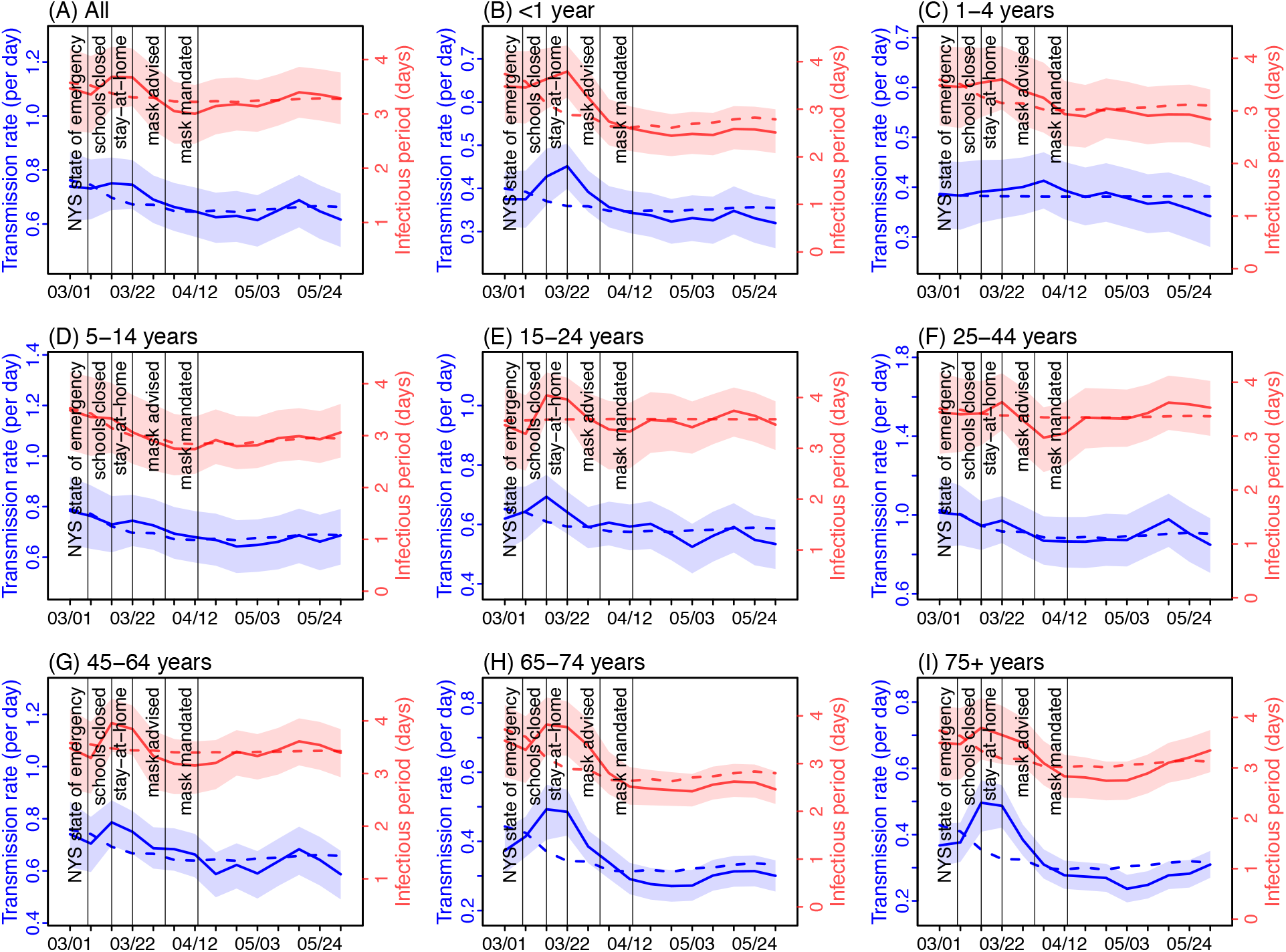
Effectiveness of face covering in reducing the transmission rate and infectious period. Solid lines show the estimated transmission rate (in blue, left y-axis) and infectious period (in red, right y-axis) using the model-inference system incorporating interventions including face covering. Surrounding areas show the 50% CrIs of model estimates. Dashed lines show corresponding estimates from a linear regression model with mobility as the sole predictor (i.e without accounting for face covering).

Given these observations, we further used the discrepancies in the two model estimates to approximate the impact of face covering on reducing COVID-19 transmission. Combining the reduction in the transmission rate and effective infectious period, we estimated that, for all ages combined, face covering contributed to a 6.6% (95% CI: 0.8 – 12.4%) reduction during the first month it was implemented and a 3.4% (95% CI: -1.9 – 8.6%) reduction over the entire 8 weeks prior to the city’s reopening (Table 1). As expected, the estimated impact varied substantially by age group. The effectiveness was 20.8% (95% CI: -0.1 – 41.6%) for 65-74 year-olds and 20.8% (95% CI: -0.9 – 42.5%) for 75+ year-olds during the first month and remained at similar levels afterwards. For 25-44 and 45-64 year-olds, two age groups with the highest infection rates (Fig 2), the effectiveness was 4.5% (95% CI: -0.6 – 9.7%) and 8.1% (95% CI: -0.1 – 16.1%) in the first month, respectively; however, it reduced substantially afterwards, likely due to reversed risk behavior. Of note, in addition to the likely lower usage rate of face covering in late May – early June, increases in risky behaviors such as large gatherings at the time^25^ may have partially obscured the effectiveness of masking.

### Retrospective projections of cases and deaths

NYC started phased reopening from the week of 6/7/2020, which allows industries to gradually reopen per a four-phase plan.^26^ For instance, manufacturing industries were allowed to reopen starting the week of 6/7/2020 (dubbed “Phase 1”), whereas real estate was allowed to reopen starting the week of 6/21/2020 (dubbed “Phase 2”), and personal care services were allowed starting the week of 7/6/2020 (dubbed “Phase 3”). As such, population mobility has increased gradually during this time, which could lead to increased transmission of unknown magnitude. Such changes also offer an opportunity to test the accuracy of our estimates – should the estimated effectiveness of reducing contact rates and utilizing face coverings be accurate, these estimates could be used to anticipate changes in transmission in response to the changing mobility and in turn the epidemic dynamics after reopening. We thus used these estimates to generate projections of cases and deaths for the 8 weeks beyond our study period, and compared the projections to available, independent corresponding observations. Overall, our projections underestimated the total number of cases (relative error of median projections: -27% over 8 weeks; Fig 5A) but were able to accurately estimate the total number of deaths (relative error: -2% over 8 weeks; Fig 5B). In addition, examination of age-grouped projections shows that the underestimation of cases was mostly among younger age groups whose case rates had increased in June (1-4, 5-14, 15-24 and 25-44 year-olds; Fig S5). These recent increases in young cases may have resulted from more young adults returning to work including some in service industries with high contact rates and, relatedly, sending their children to childcare and/or summer camps due to a lack of caretakers at home [information from NYC Department of Health and Mental Hygiene (DOHMH) community investigation; unpublished]. In addition, increased risk behaviors of some young individuals (e.g., large parties without physical distancing^25^) may have also contributed to the increased cases among young adults in late June.^27^ Consistently, COVID-19 associated mortality, mostly occurring among older adults continued to decrease and were accurately predicted for different age groups (Fig S6).

**Fig 5.**
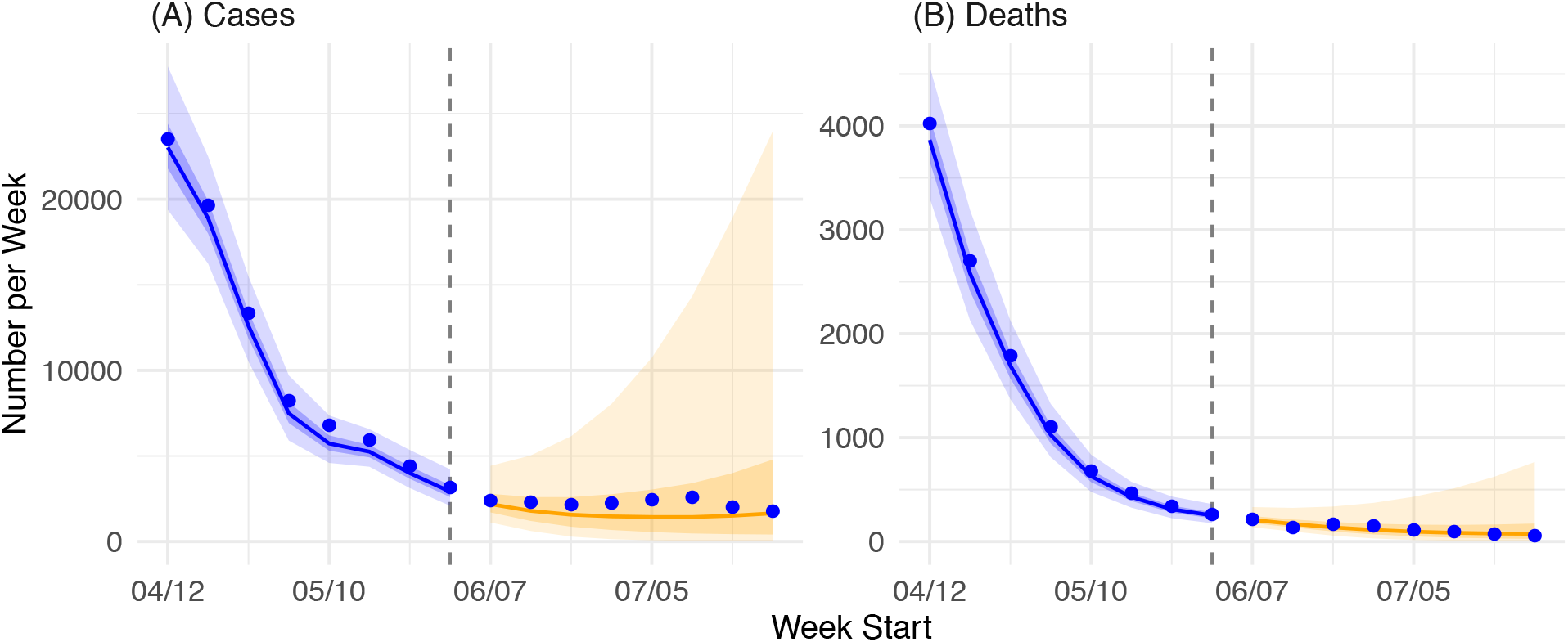
Projections of COVID-19 cases and deaths eight weeks beyond the study period. Blue 767 dots show confirmed cases by week of diagnosis and deaths by week of death, as observed 768 by the surveillance system. Blue lines show model median estimates; surrounding shades 769 show 50% and 90% CrIs. Orange lines show model projected median weekly cases and 770 deaths; surrounding shades show 50% and 90% CIs of the projection.

## Discussion

The spring 2020 pandemic wave in NYC, the first epidemic center in the US, provides a test case to study COVID-19 epidemiological characteristics and the effectiveness of public health interventions. Through comprehensive modeling, we have reconstructed the transmission dynamics and estimated the effectiveness of two major interventions, social distancing and mandatory face covering in public. Our results show that reducing contact rates (mainly via school closures and voluntary or mandated stay-at-home measures) likely contributed to the largest reduction in transmission in the population overall (∼70%) and for most age groups (>50% for all age groups). Widespread use of face covering likely contributed to an additional ∼7% overall reduction and up to ∼20% reduction among 65+ year-olds during the first month face covering was mandated in public places. Our findings largely consolidate previous model estimates on the impact of lockdown-like measures^4,28,29^ and studies on face covering in reducing COVID-19 transmission. These findings provide insights that can inform COVID-19 mitigation efforts in the coming months before the majority of population can be protected through mass-vaccination, as well as control strategies for other (re)emerging infections in the future.

Lockdown-like measures where confinement at home is encouraged or mandated through school closures, telework policies, closure of non-essential businesses, and stay-at-home orders have been a major control measure to curb COVID-19 spread. In effect, such measures reduce population contact rates and thus transmission. Previous modeling studies estimated that lockdowns reduced COVID-19 transmission (measured by *Rt*) by 58% in Wuhan, China,^4^ 45% (95% CI: 42-49%) in Italy,^29^ and 77% (95% CI: 76-78%) in France.^28^ Our estimate for NYC overall (∼70%) is consistent with these previous estimates. In addition, our estimates show that reducing population contact rates effectively reduced transmission across all age groups (ranging from a 51% reduction among 1-4 year-olds to 83% among 5-14 year-olds; Table 1). Together, these findings underscore the importance of reducing contact rates through, for example, physical distancing in places with continuous community transmission of COVID-19.

The use of surgical masks or cloth face coverings has been another major preventive measure for COVID-19. Studies overall have shown that surgical masks could substantially reduce onward transmission albeit with a large range of efficacy estimates across settings.^30^ However, it remains unclear the overall effectiveness of universal face covering requirements at the population level, especially during a pandemic, due to several factors: 1) The overall effectiveness depends on compliance which may vary across subpopulations and time; 2) Improper use of face coverings (e.g. without covering the nose and/or mouth or improper handling^31^) can reduce the effectiveness of face covering; 3) Face coverings are required and mostly worn in public and thus likely have a lower impact in private settings, particularly in reducing household transmission; consequently, the relative impact of face covering depends on the relative contribution of different sources of transmission (e.g. household vs. community) at a given time and *vice versa*; and 4) Use of face coverings may lead to complacency and less stringent adherence to social distancing and stay-at-home behaviors. Here we estimated a ∼7% reduction in overall transmission during the first month of the face covering mandate. However, the estimated effectiveness varied largely across age groups with much higher effectiveness among older adults (∼20% for both 65-74 and 75+ year-olds vs. <10% for other age groups). This discrepancy was likely due to the differential compliance and types of face covering used. Observational studies in Wisconsin and surveys nationwide in April/May reported about 2-fold higher rates of face covering usage among older adults versus younger adults and minors.^22-24^ In addition, due to the shortage of surgical masks during March–May,^32^ older adults at higher risk of severe COVID-19 infection were more likely to use surgical masks whereas younger age groups more frequently used non-medical cloth coverings, which are often less effective^33,34^ (e.g. measured ultrafine filtration efficiency is ∼50% for surgical masks vs. ∼10-25% for T-shirt and ∼25-35% for cotton covers^34^).

When lockdown-like measures are lifted, residents will spend more time outside their homes than during the lockdown. Adjusting for the time spent outside of homes (∼8.3 hours in April 2020 vs. ∼11.5 hours in June-July 2020 and ∼13.5 hours pre-pandemic; NYC data^35^), universal face covering would have reduced overall transmission by ∼9–11% (i.e., 6.6% multiplied by a factor of 1.4–1.6) during reopening, given the same rates of face covering as in April. However, if the same effectiveness among older adults were achieved among other age groups, universal face covering could reduce overall transmission by up to ∼28–32% (i.e., 20% multiplied by a factor of 1.4–1.6). The implication of this latter estimate is two-fold. On the one hand, it suggests that for places with high level of transmission, implementing face covering *alone* is likely insufficient to lower the effective reproductive number *R*_*t*_ to <1 in order to control the epidemic [for instance, for a city with an *R*_*0*_ = 3, the resulting *R*_*t*_ would be 3 × (1– 30%) = 2.1]. This finding is consistent with the observed resurgence of COVID-19 cases in NYC during fall/winter 2020 despite the concurrent high usage rate of face coverings (∼90% of survey respondents in NYC reported always or frequently wearing masks in public in July 2020;^36^ and this number was likely higher during fall/winter 2020). On the other hand, our findings also suggest that improving effective usage rates of face coverings, especially among younger age groups, could significantly mitigate the risk of resurgence of COVID-19 infections during re-opening (i.e., ∼30% reduction without compromising economic growth). It is thus crucial for future research to understand reasons for the use/non-use and selection of face coverings by age group to inform strategies to increase consistent and correct mask use in settings where social distancing is not possible.

It is important to note, however, that not all individuals have the same opportunities to physically distance and/or adopt face coverings during a pandemic, despite government mandates. For instance, over one million frontline workers in NYC (e.g., healthcare workers, transportation workers, janitors, and grocery clerks, which comprise 25 percent of the city’s workforce) had to continue their essential work during the pandemic.^37^ In addition, data from the U.S. Bureau of Labor Statistics suggest Black and Latino communities have less opportunities to work from home.^38^ Consequently, NYC neighborhoods with more frontline workers and/or Black and Latino residents tended to have lower reductions in population mobility during the pandemic.^35^ In NYC, these communities also experience a number of social conditions that are thought to exacerbate COVID-19, including overcrowded multigenerational households, poverty, and high prevalence of chronic diseases. These communities, known to also carry a higher burden of underlying health conditions, suffered greater impacts from COVID-19 and have expressed fear and experiences of racialized bias when wearing a face covering.^39,40^ Further research is warranted to investigate such health disparities. In addition, future policies should take into account structural inequities in labor trends, overcrowded housing, and underlying conditions and adopt additional preventive measures to protect those vulnerable communities.

We also note there remain large uncertainties in our estimates due to several limitations. First, we used population mobility as a proxy for contact rates rather than more direct measures. Similar approximation and uncertainty applied to our estimates of the effectiveness of face covering. Future studies are thus warranted for further assessment. For instance, large population scale surveys documenting changes of the intensity and pattern of contact during the pandemic could provide more accurate measures of contact rates among different age groups and over time. Second, while we restricted our analysis on the effectiveness of face covering to a period when masks were mandated, there remain other residual confounding effects. For instance, increased awareness of COVID-19 and health risk among key age groups such as the elderly may have contributed to further reductions of transmission through other precautions in addition to face covering; this may have led to an overestimation of the effectiveness of face covering for those age groups. Third, here we focused on estimating the effectiveness of interventions in the general population without segregating key settings with intense transmission (e.g., long-term care facilities). Future studies should assess the impact of interventions targeting such high-risk settings. Lastly, our estimates here were largely based on the first wave of the pandemic and may not fully capture subsequent changes in awareness and perception of COVID-19 and related behavioral adjustment during later waves. However, we have also used a similar methodology to estimate the effectiveness of reducing contact rates and face covering under different city reopening schedules and generated long-term projections for NYC; the projections generated during June 2020 for the period of June 2020 – May 2021 have been consistent with observations up to the end of 2020 (i.e., at the time of this writing; see the projected resurgence and second wave in Yang et al.^41^ and comparison with available data in Fig S7). These results thus support the robustness of our estimates here.

Our study also has several strengths. In particular, our estimates were based on comprehensive model-inference incorporating multiple data streams and further evaluated using model projections. Our results thus provide an assessment of two major public heath interventions (reducing contact rates and face covering) at the population level where the overall effectiveness depends on multiple factors in addition to the efficacy of a given intervention. Altogether, our estimates support the need for multiple interventions (including reducing contact rates by, e.g., restricting occupancy, universal face covering, and, albeit not studied here, testing, contact tracing, isolation and timely treatment of cases) in order to effectively mitigate the spread of COVID-19 as it continues to pose threats to public health.

## Methods

### Data

COVID-19 cases included all laboratory-confirmed cases by week of diagnosis reported to the NYC DOHMH. Mortality data by week of death combined confirmed and probable COVID-19-associated deaths. Confirmed COVID-19-associated deaths were defined as those occurring in persons with laboratory-confirmed SARS-CoV-2 infection, and probable COVID-19 deaths were defined as those with COVID-19, SARS-CoV-2, or a similar term listed on the death certificate as an immediate, underlying, or contributing cause of death, but did not have laboratory-confirmation of COVID-19.^42^ For this study, both weekly case and mortality data were aggregated by age group (<1, 1-4, 5-14, 15-24, 25-44, 45-64, 65-74, and 75+ years) for each of the 42 United Hospital Fund (UHF) neighborhoods,^43^ according to the patient’s residential address. All data were retrieved on Sep 4, 2020. For a summary of the spatial variations across the 42 neighborhoods, see Table S3 in the Appendix of Yang et al.^12^

The mobility data, used to model changes in population contact rates due to public health interventions implemented during the pandemic (e.g., social distancing), came from SafeGraph^35,44^ and contained counts of visitors to locations in each zip code from the same zip code and others, separately, based on mobile device locations. The released data were anonymized and aggregated in weekly intervals. We spatially aggregated these data to the UHF neighborhood level, for both intra and inter UHF neighborhood mobility. In addition, SafeGraph also provided an aggregate measure of the length of time spent outside of the home during each week.

This study was classified as public health surveillance and exempt from ethical review and informed consent by the Institutional Review Boards of both Columbia University and NYC DOHMH.

### Network transmission model

The epidemic model used in this study was described in Yang et al. 2020^11,12^ Briefly, the model simulated intra- and inter neighborhood transmission of COVID-19 using a susceptible-exposed-infectious-removed (SEIR) network model:

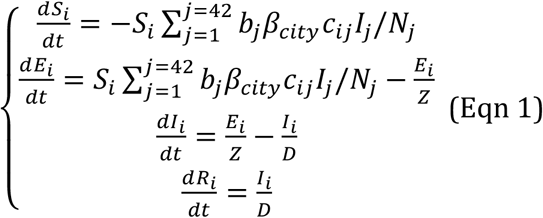

where *S*_*i*_, *E*_*i*_, *I*_*i*_, *R*_*i*_, and *N*_*i*_ are the numbers of susceptible, exposed (but not yet infectious), infectious, and removed (either recovered or deceased) individuals and the total population, respectively, from a given age group in neighborhood *i*. Note that due to model complexity and a lack of information for parameterizing interactions among age groups, we modeled each age group separately (i.e., combining all sources of transmission to each age group; see further detail on parameter estimation below); as such, Eqn 1 describes the spatial transmission across neighborhoods without interactions among age groups. *β* _*city*_ is the citywide transmission rate, which incorporated seasonal variation as observed for OC43, a beta-coronavirus in humans from the same genus as SARS-CoV-2.^12^ To allow differential transmission in each neighborhood, we included a multiplicative factor, *b*_*i*_, to scale neighborhood local transmission rates. *Z* and *D* are the latency and infectious periods, respectively (Table S4).

The contact rates (*c*_*ij*_) in each neighborhood over time and connectivity among neighborhoods were computed based on mobility data. The model also accounted for delays from infection to diagnosis using two parameters (gamma distribution with mean *T*_*d*_ and standard deviation *T*_*sd*_ estimated along with other parameters) and death (based on observed time from diagnosis to death) as well as infection detection rate using a parameter *r* (estimated along with other parameters). For further detail, please refer to Yang et al. 2020.^11,12^

### Parameter estimation

To estimate model parameters (e.g., *b*_*i*_, *β*_*city*_, *Z, D, r*, and *infection fatality risk*, for *i*=1,…,42) and state variables (e.g., number of susceptible and infectious individuals in each neighborhood) for each week, we ran the network-model stochastically with a daily time step in conjunction with the ensemble adjustment Kalman filter (EAKF)^45^ and fit to weekly case and mortality data from the week starting March 1 to the week ending June 6, 2020. The posterior distribution of each model parameter/variable was updated for that week at the same time.^45^ This parameter estimation process was done separately for each of the eight age groups (i.e. <1, 1-4, 5-14, 15-24, 25-44, 45-64, 65-74, and 75+ years). To include transmission from other age groups, we used measured intra and inter-group contacts from the POLYMOD study^21^ to compute the total number of contacts made with each age group and adjusted the *prior* range of the transmission rate (*β*_*city*_) for each age group accordingly. The posterior estimate was computed based on cases and mortality data for each group, which included all sources of infection. Thus, the estimated transmission rate for each age group nevertheless included all sources of transmission. To account for stochasticity in model initiation, we ran the parameter estimation process independently 10 times. Results for each age group were combined from these 10 runs (each with 500 model realizations). We computed age-specific *R*_*t*_, the effective reproductive number during week-*t*, from the posterior estimates of transmission rate (*β*_*city*_ and *b*_*i*_), infectious period (*D*), contact matrix (*c*_*ij*_), susceptibility and population size in the neighborhood using the next generation method.^46^ We computed *R*_*t*_, *β*_*city*_, and *D* estimates for all ages overall as a weighted average of the age-specific estimates with weights equal to the population fraction in each age group.

### Estimating the effectiveness of reducing contact rates

The *R*_*t*_ estimates from the model-inference system capture changes in transmission due to various interventions, i.e., the overall effectiveness of all implemented interventions. To separately estimate the effectiveness of interventions that reduce contact rates, we used human mobility as a measure of population contact rate to estimate the changes in *R*_*t*_ in response to changing population contact rates. Specifically, we regressed the *R*_*t*_ estimates from the full model-inference system on the mobility data: *R*_*t*_ = *a*_0_ + *a*_1_ *M*_*ave,t*_ (Eqn 2), where *M*_*ave,t*_ is the mean of all intra-neighborhood mobility at week-*t*. We then computed the effectiveness of reducing contact rate based on 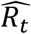, the *R*_*t*_ estimate from this regression model solely based on the observed mobility. That is, the reduction in *R*_*t*_ by week-*t*, likely due to reducing contact rate, was computed as 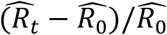 (Eqn 3). To test the robustness of our method, we performed the same analysis for individual UHF neighborhoods (*n* = 42).

### Estimating the effectiveness of face covering/masking

In addition to changes in population contact rates, face covering/masking was another major control measure implemented beginning the week of April 12, 2020 when NYC mandated residents wear face coverings in public places. To estimate the effectiveness of face covering, we first estimated the changes in transmission rate and effective infectious period, two model parameters determining *R*_*t*_, due to changes in mobility (as opposed to masking) using regression models similar to Eqn 2. Specifically, we regressed the estimated citywide transmission rate (or effective infectious period) from the full model-inference system on average mobility: *Y*_*t*_ = *a*_0_ + *a*_1:_ *M*_*ave,t*_ (Eqn 4), where *Y*_*t*_ is *β*_*city*_ or *D*. We then computed the relative reduction in transmission rate (or effective infectious period) due to face covering as 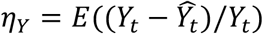, where E(·) gives the mean over the relevant timeframe (here we estimated two timeframes, i.e., 1 month following the mandate and over 8 weeks up to 6/6/2020). Combining both reductions, we computed the effectiveness of face covering as *η* = 1 − (1 − *η*_*β*_)(1 − *η*_*D*_). Of note, while mechanistically face coverings act primarily by reducing the probability of transmission (i.e., transmission rate), here we included both the potential impact on the transmission rate (*η*_*β*_) and effective infectious period (*η*_*D*_), mainly because the multiplicative relationship of the two variables with *R*_*t*_ makes it challenging to separate the two effects. Nevertheless, reductions in the infectious period via face covering are possible. A recent study on animals showed that masking could reduce the severity of infection;^47^ if persons with milder infection experience shorter duration of viral shedding (there is some evidence for this, e.g., from He et al.^48^), milder symptoms in individuals infected while wearing face covering could lead to shorter infectious period of these individuals.

### Projections of cases and deaths

To evaluate the accuracy of model estimates, in particular, the effectiveness of transmission reduction by reducing contact rates and use of face covering, we tested if these estimates along with the model could generate accurate predictions of cases and deaths for 8 weeks beyond the study period (i.e. from the week of 6/7/2020 to the week of 7/26/2020). We first projected the citywide transmission rate and infectious period based on observed mobility using Eqn 4; these estimates thus accounted for changes due to changes in contact rates. To incorporate the reduction in transmission by face covering, we further reduced the projected city transmission rate by a factor of 1 − *η*_*β*_*p*_*out*_ and the infectious period by a factor of 1 − *η*_*D*_*p*_*out*_ where *p*_*out*_ is a factor to adjust for time spent outside of the home during each week. To reflect longer-term usage rates of face covering, we used *η*_*β*_ and *η*_*D*_ estimated during the entire 8 weeks face covering was required (i.e. from the week of 4/12/2020 to the week of 5/31/2020). Finally, we used estimates of population susceptibility and infection rates at the end of the week of 5/31/2020 to model initial conditions and integrated the SEIR network model forward stochastically for 8 weeks using the projected transmission rate and infectious period.

## Data Availability

The COVID-19 case and mortality data used here are permitted via a data use and nondisclosure agreement with the NYC DOHMH. Part of the data are publicly available at https://github.com/nychealth/coronavirus-data. For full access, please contact the NYC DOHMH.

## Acknowledgments

This study was supported by the National Institute of Allergy and Infectious Diseases (AI145883), the National Science Foundation Rapid Response Research Program (RAPID; 2027369), and the NYC DOHMH. We thank the NYC DOHMH Bureau of Vital Statistics team and EpiData team; in particular, Mary Huynh, Sharon K. Greene, Alice Yeung, Anne Fine, Miranda S. Moore and Kevin Guerra, for data management and provision. We thank Jennifer Brite at NYC DOHMH for coordinating discussions on modeling COVID-19 in NYC and Alana Tornello for amplifying community voices. We also thank Columbia University Mailman School of Public Health for high performance computing, Safe Graph (safegraph.com) for providing the mobility data, and Sasikiran Kandula at Columbia University for compiling the mobility data used in this study. We are also grateful to Sasikiran Kandula, Sharon K. Greene, Anne Fine, and Hannah Helmy for helpful feedback on this manuscript.

## Author contributions

WY designed study, conducted the modeling analyses, and wrote the first draft. J Shaff provided inputs on COVID-19 transmission dynamics, public health interventions and community responses in NYC and critically revised the manuscript. J Shaman contributed to study design, interpreted model results, and critically revised the manuscript.

## Supplemental Tables and Figures

### Supplemental Tables

**Table S1.** Total number of reported cases and estimated cumulative infection rate by the week of 5/31/20. Case rate was computed as the number of total cases divided by the population size of the corresponding age group. Estimated infection rate was estimated by model-inference system, normalized to the corresponding population size; numbers are median (and 95% CrIs).

| Age | Total number of cases | Case rate (%) | Estimated infection rate (%) |
| --- | --- | --- | --- |
| all | 205693 | 2.45 | 17.6 (13.2, 25.5) |
| <1 | 515 | 0.46 | 4.4 (2.9, 10.5) |
| 1-4 | 609 | 0.14 | 1.4 (0.9, 3.7) |
| 5-14 | 2646 | 0.28 | 10.5 (6.5, 24.5) |
| 15-24 | 12562 | 1.28 | 12.0 (8.4, 19.5) |
| 25-44 | 64764 | 2.45 | 22.7 (16.8, 31.5) |
| 45-64 | 74833 | 3.64 | 23.1 (18.4, 29.6) |
| 65-74 | 25469 | 3.64 | 15.2 (11.4, 21.7) |
| 75+ | 24295 | 4.44 | 12.7 (9.8, 18.4) |

**Table S2.** Estimated reproductive number by week and age group. Numbers are median (and interquartile range) of the posterior estimates.

| date | Age Groups |  |  |  |  |  |  |  |  |
| --- | --- | --- | --- | --- | --- | --- | --- | --- | --- |
|  | all | <1 | 1-4 | 5-14 | 15-24 | 25-44 | 45-64 | 65-74 | 75+ |
| 3/1/20 | 2.99 (2.32, 3.86) | 1.52 (1.19, 1.96) | 1.56 (1.22, 2.02) | 3.15 (2.47, 4.09) | 2.47 (1.93, 3.22) | 4 (3.1, 5.15) | 2.97 (2.3, 3.86) | 1.77 (1.37, 2.29) | 1.75 (1.35, 2.27) |
| 3/8/20 | 2.25 (1.64, 2.98) | 1.43 (1.11, 1.85) | 1.28 (1, 1.7) | 2.35 (1.74, 3.18) | 2.02 (1.53, 2.64) | 2.46 (1.57, 3.48) | 2.49 (1.97, 3.12) | 1.98 (1.55, 2.44) | 1.84 (1.48, 2.29) |
| 3/15/20 | 2.14 (1.62, 2.74) | 1.63 (1.32, 2) | 1.17 (0.92, 1.48) | 1.52 (1.13, 2.06) | 2.38 (1.86, 3) | 1.45 (0.97, 2.03) | 2.99 (2.3, 3.68) | 2.84 (2.33, 3.51) | 2.87 (2.33, 3.52) |
| 3/22/20 | 1.37 (1.08, 1.68) | 1.59 (1.35, 1.89) | 1.05 (0.85, 1.29) | 0.99 (0.74, 1.3) | 1.29 (1.03, 1.57) | 1 (0.76, 1.26) | 1.37 (1.06, 1.71) | 2.58 (2.15, 3.04) | 2.52 (2.06, 3.02) |
| 3/29/20 | 0.93 (0.73, 1.13) | 1.22 (0.97, 1.44) | 1.06 (0.82, 1.3) | 0.88 (0.66, 1.14) | 0.96 (0.75, 1.15) | 0.7 (0.52, 0.88) | 0.89 (0.71, 1.06) | 1.43 (1.17, 1.65) | 1.46 (1.27, 1.66) |
| 4/5/20 | 0.63 (0.5, 0.75) | 0.74 (0.6, 0.88) | 0.84 (0.65, 1.03) | 0.53 (0.41, 0.68) | 0.77 (0.59, 0.91) | 0.44 (0.33, 0.56) | 0.62 (0.5, 0.73) | 0.9 (0.79, 1.02) | 0.9 (0.78, 1.02) |
| 4/12/20 | 0.56 (0.45, 0.67) | 0.67 (0.54, 0.79) | 0.69 (0.53, 0.84) | 0.5 (0.38, 0.62) | 0.72 (0.57, 0.83) | 0.44 (0.34, 0.55) | 0.57 (0.47, 0.67) | 0.63 (0.55, 0.71) | 0.65 (0.57, 0.74) |
| 4/19/20 | 0.59 (0.48, 0.7) | 0.67 (0.54, 0.8) | 0.68 (0.54, 0.81) | 0.57 (0.44, 0.7) | 0.81 (0.66, 0.93) | 0.55 (0.42, 0.68) | 0.49 (0.41, 0.57) | 0.6 (0.52, 0.68) | 0.66 (0.58, 0.74) |
| 4/26/20 | 0.62 (0.5, 0.73) | 0.62 (0.5, 0.75) | 0.77 (0.62, 0.91) | 0.54 (0.42, 0.67) | 0.75 (0.61, 0.86) | 0.58 (0.45, 0.72) | 0.6 (0.5, 0.71) | 0.58 (0.5, 0.67) | 0.63 (0.56, 0.71) |
| 5/3/20 | 0.66 (0.54, 0.77) | 0.7 (0.56, 0.84) | 0.78 (0.62, 0.91) | 0.56 (0.44, 0.67) | 0.72 (0.61, 0.8) | 0.68 (0.54, 0.82) | 0.68 (0.56, 0.78) | 0.6 (0.52, 0.67) | 0.55 (0.48, 0.63) |
| 5/10/20 | 0.82 (0.67, 0.95) | 0.71 (0.56, 0.86) | 0.75 (0.6, 0.88) | 0.6 (0.47, 0.73) | 0.88 (0.73, 0.98) | 0.9 (0.72, 1.06) | 0.88 (0.74, 0.99) | 0.73 (0.64, 0.81) | 0.65 (0.57, 0.74) |
| 5/17/20 | 1.03 (0.83, 1.18) | 0.82 (0.65, 0.99) | 0.8 (0.65, 0.93) | 0.73 (0.57, 0.88) | 1.1 (0.89, 1.24) | 1.21 (0.95, 1.42) | 1.1 (0.91, 1.24) | 0.78 (0.69, 0.87) | 0.81 (0.72, 0.92) |
| 5/24/20 | 0.89 (0.73, 1.03) | 0.78 (0.61, 0.95) | 0.79 (0.63, 0.91) | 0.69 (0.53, 0.83) | 0.94 (0.78, 1.06) | 0.96 (0.76, 1.13) | 0.95 (0.78, 1.08) | 0.79 (0.7, 0.88) | 0.89 (0.78, 0.99) |
| 5/31/20 | 0.79 (0.66, 0.91) | 0.71 (0.55, 0.87) | 0.71 (0.56, 0.83) | 0.7 (0.55, 0.86) | 0.85 (0.72, 0.95) | 0.79 (0.64, 0.93) | 0.77 (0.65, 0.89) | 0.72 (0.64, 0.79) | 1.04 (0.93, 1.15) |

**Table S3.** Correlation of key epidemiological parameters with population mobility during the week of March 1 – the week of May 31, 2020.

| Age | Rt (ignore<br>susceptibility) | Rt | Transmission<br>rate | Infectious period |
| --- | --- | --- | --- | --- |
| all | 0.96 | 0.96 | 0.73 | 0.54 |
| <1 | 0.76 | 0.77 | 0.41 | 0.70 |
| 1-4 | 0.93 | 0.93 | 0.05 | 0.65 |
| 5-14 | 0.99 | 0.99 | 0.86 | 0.92 |
| 15-24 | 0.90 | 0.91 | 0.52 | -0.05 |
| 25-44 | 0.93 | 0.96 | 0.80 | 0.27 |
| 45-64 | 0.88 | 0.90 | 0.60 | 0.24 |
| 65-74 | 0.57 | 0.63 | 0.54 | 0.65 |
| 75+ | 0.55 | 0.60 | 0.50 | 0.64 |

**Table S4.**
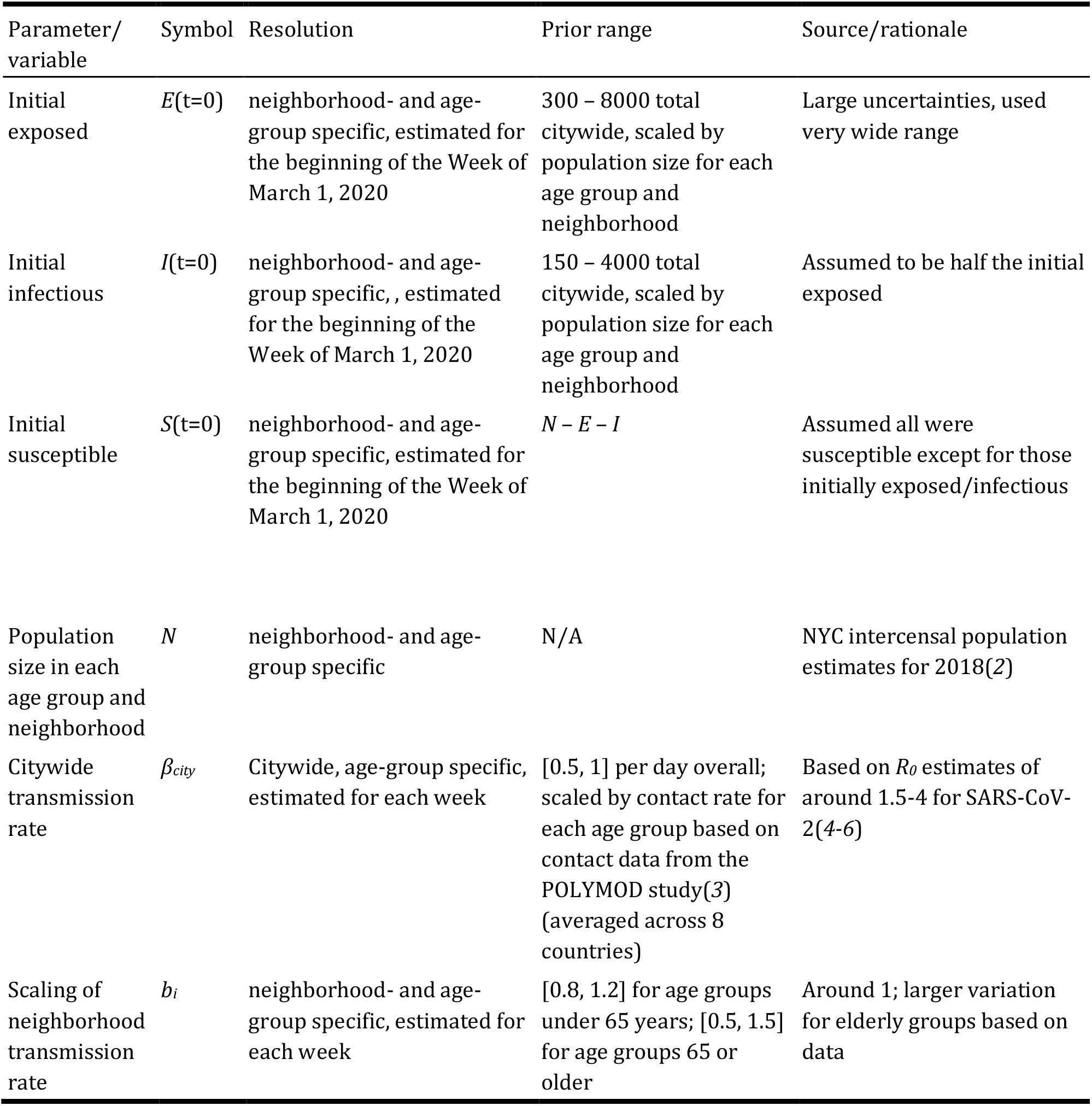
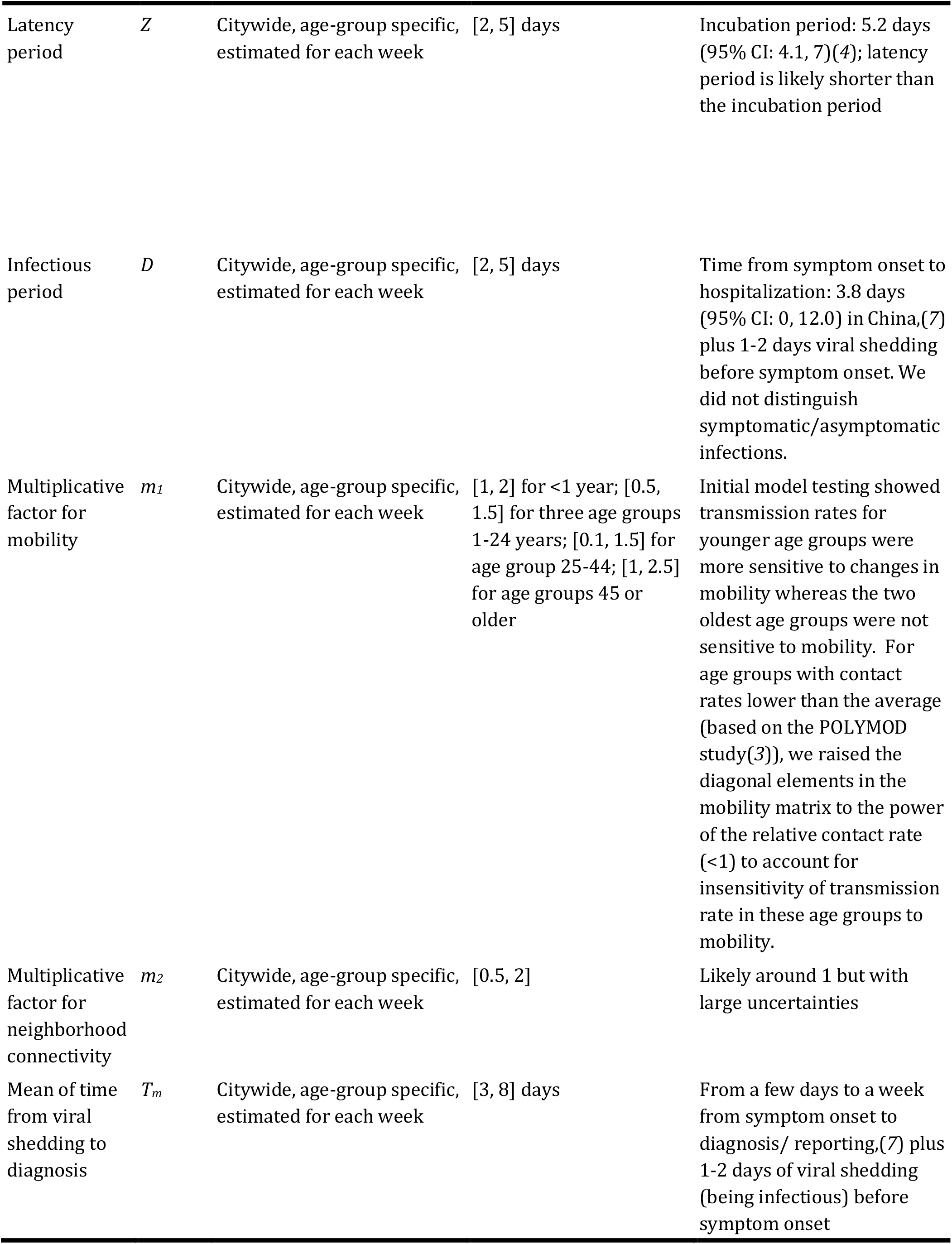
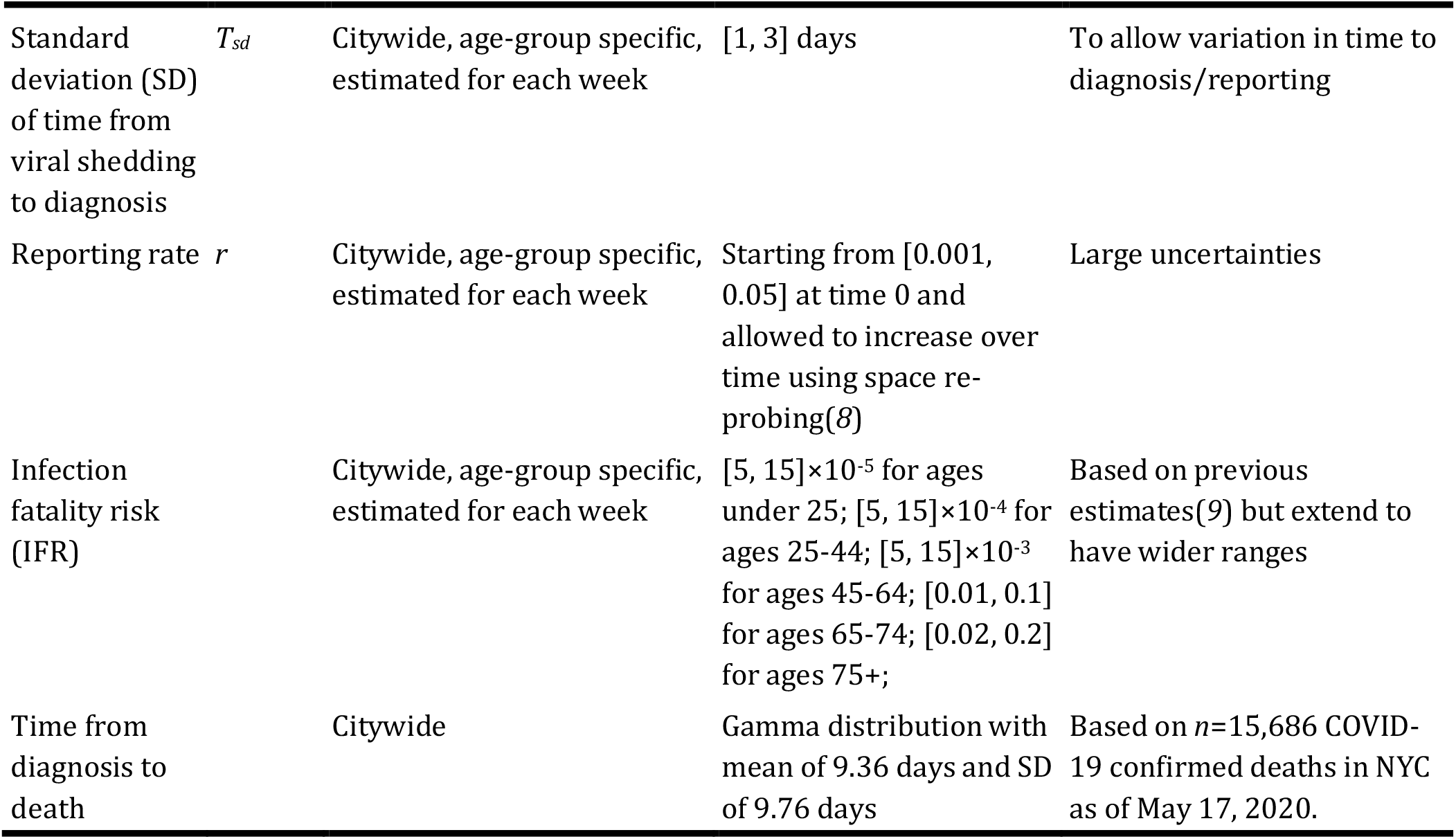
Prior ranges for main model parameters and variables. The spatial, temporal, and age resolution of each parameter or variable, estimated in the model-inference system, is specified in the column “Resolution”. Note posterior parameter estimates can extend outside the specified prior ranges. Note this is the same as Table S1 in Yang et al.(*1*)

## Supplemental Figures

**Fig S1.**
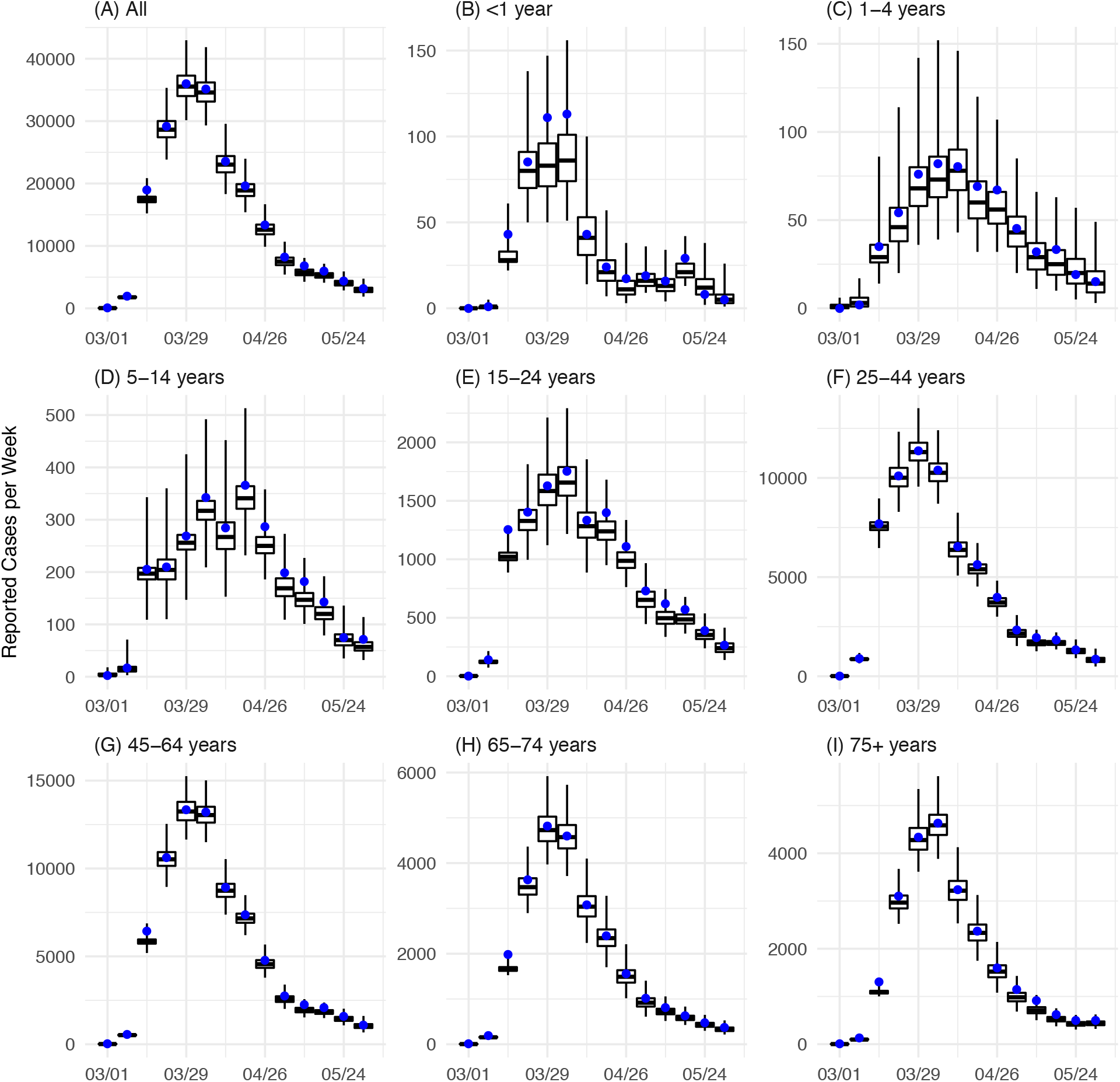
Model fits of reported confirmed COVID-19 cases. Blue dots show reported number of cases by age group and week of diagnosis. Boxes show the model fitted weekly number of cases by age group. Box edges, thick middle lines, and whiskers show the 2.5^th^, 25^th^, 50^th^, 75^th^, and 97.5^th^ percentiles of model estimates.

**Fig S2.**
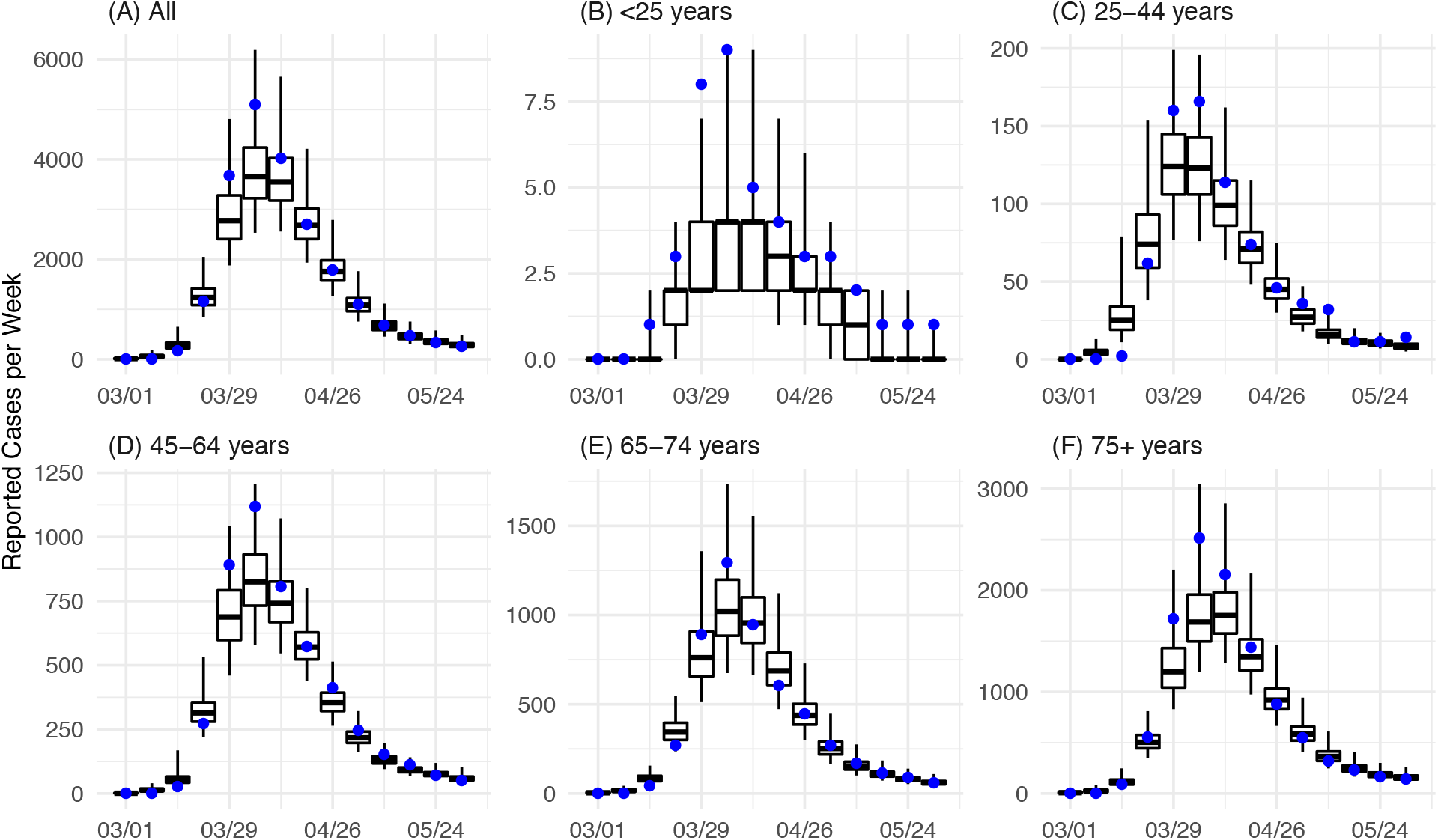
Model fits of reported COVID-19 associated deaths. Blue dots show reported weekly number of deaths by age group. Boxes show the model fitted weekly number of deaths by age group. Box edges, thick middle lines, and whiskers show the 2.5^th^, 25^th^, 50^th^, 75^th^, and 97.5^th^ percentiles of model estimates. Note that deaths among <25 year-olds are combined due to low counts.

**Fig S3.**
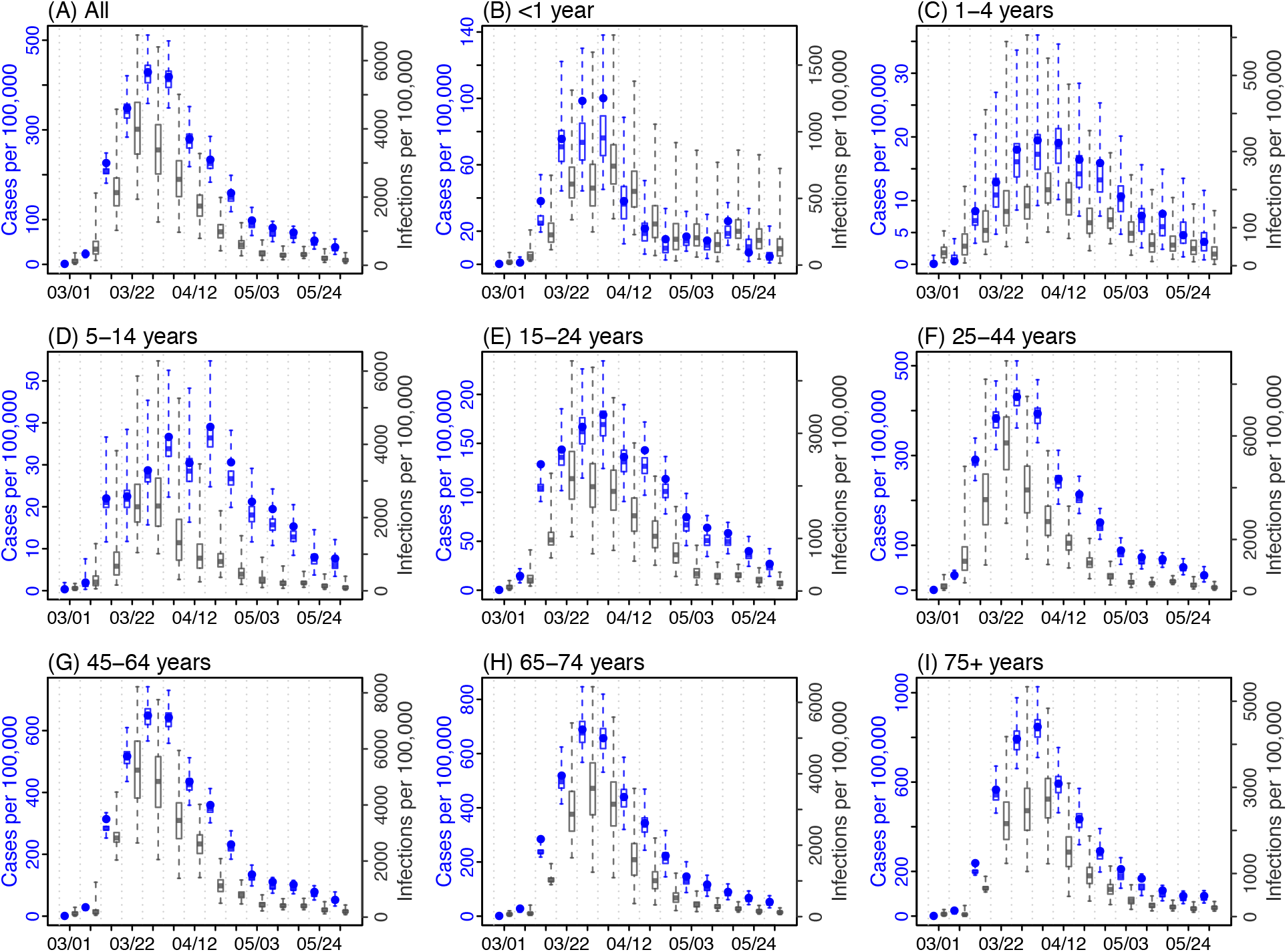
Estimated case rates and infection rates by age group. Blue dots show confirmed case rates by age group and week of diagnosis. Blue boxes (left y-axis) show the model fitted weekly case rates and grey boxes (right y-axis) show the model estimated weekly infection rates by age group. Box edges, thick middle lines, and whiskers show the 2.5^th^, 25^th^, 50^th^, 75^th^, and 97.5^th^ percentiles of model estimates.

**Fig S4.**
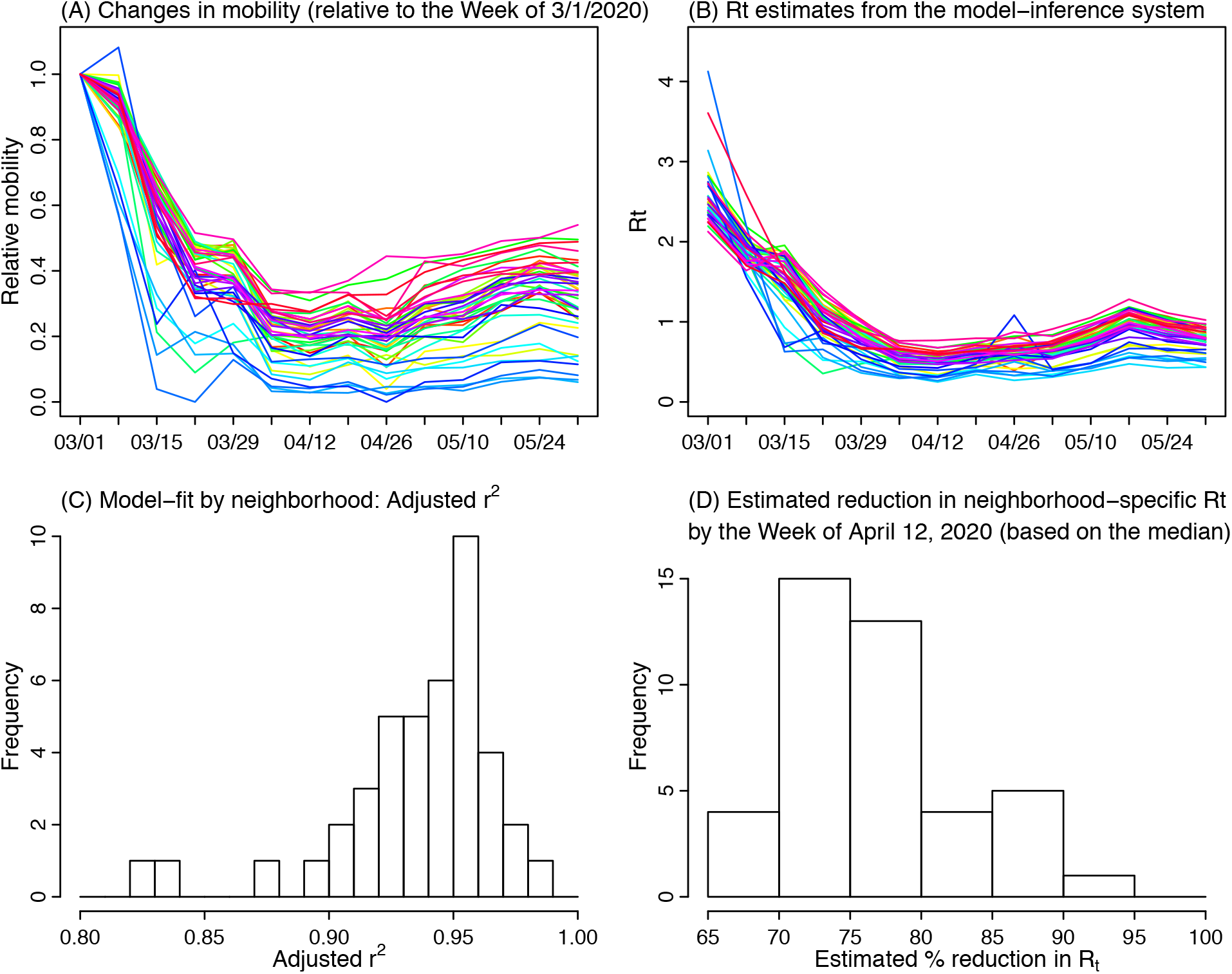
Sensitivity analysis on the effectiveness of reducing contact rate by neighborhood. There are 42 United Hospital Fund (UHF) neighborhoods in NYC. (A) shows the changes in human mobility during the pandemic by neighborhood (each colored line). The reductions were substantial in all neighborhoods but to varying degrees. (B) shows the estimated *R*_*t*_ (combining all ages) for each week and neighborhood (each colored line). Note these estimates did not account for changes in susceptibility so as to restrict to changes due to interventions. (C) shows the adjusted r^2^ of linear regression model fitting the mobility data (A) to the *R*_*t*_ estimates (B), for each neighborhood, per Eqn 2 in the main text. Adjusted r^2^ for most neighborhoods was >0.9. (D) shows the estimated reduction in R_t_ by the Week of April 12, 2020 based on the Eqn 3 in the main text, for each neighborhood. The histogram is based on the median estimated reduction in R_t_.

**Fig S5.**
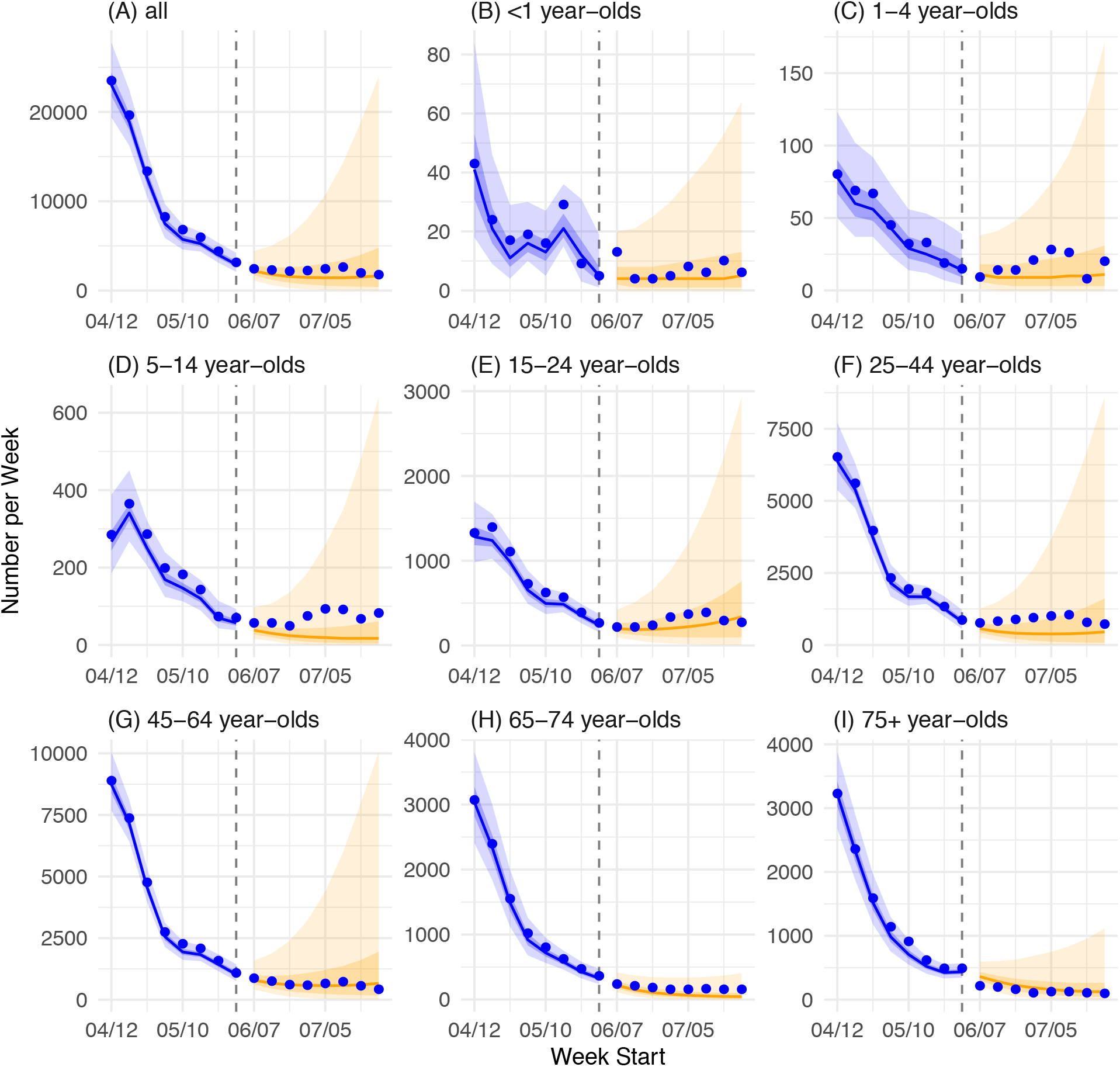
Projections of COVID-19 cases by age group eight weeks beyond the study period. Blue dots show observed confirmed cases by week of diagnosis (those after the Week of 5/31/2020 were not used in the model). Blue lines show model median estimates; surrounding shades show 50% and 90% CrIs. Orange lines show model projected median weekly cases and deaths; surrounding shades show 50% and 90% CIs of the projection.

**Fig S6.**
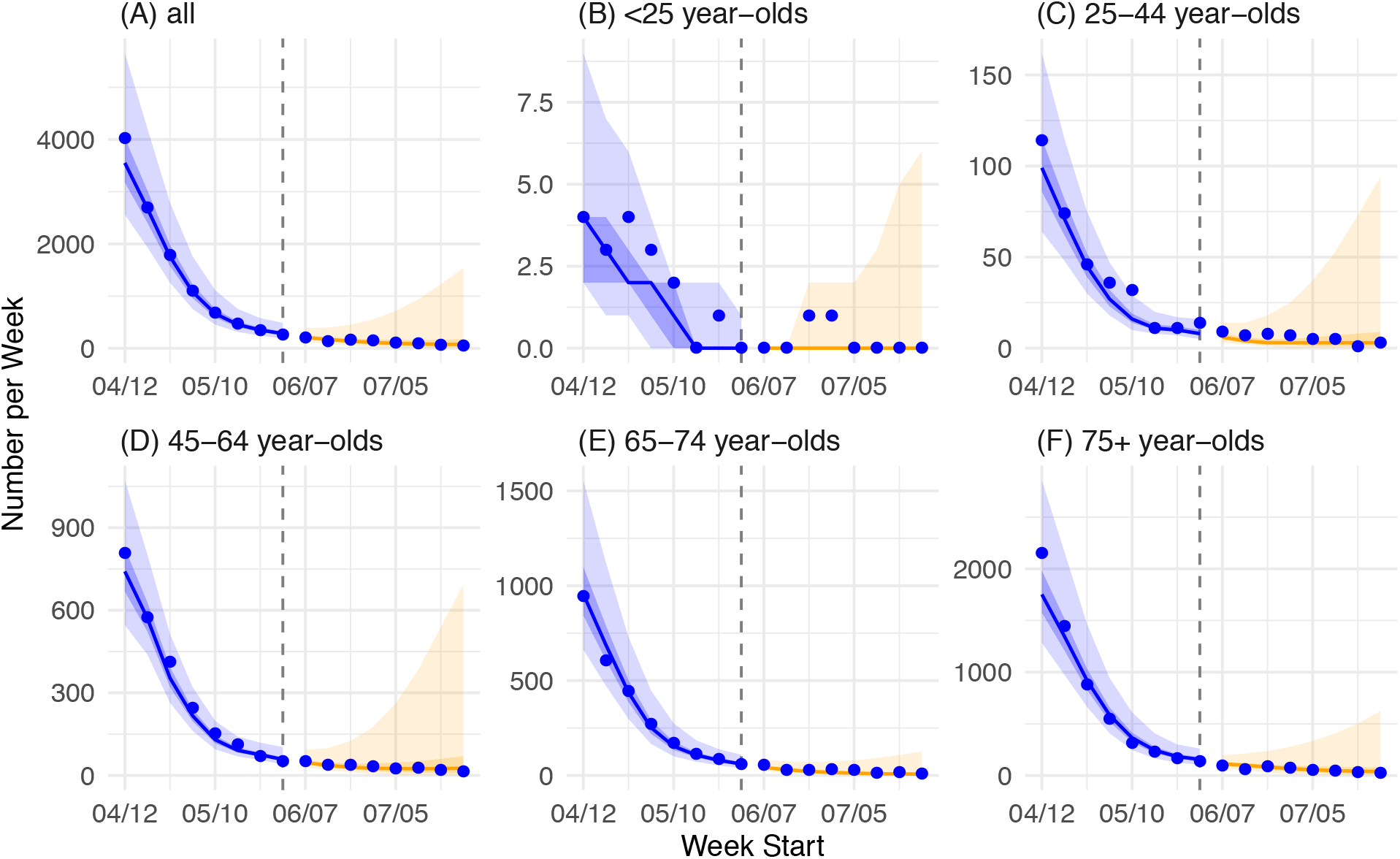
Projections of COVID-19 associated deaths by age group eight weeks beyond the study period. Blue dots show observed weekly deaths (those after the Week of 5/31/2020 were not used in the model). Blue lines show model median estimates; surrounding shades show 50% and 90% CrIs. Orange lines show model projected median weekly cases and deaths; surrounding shades show 50% and 90% CIs of the projection.

**Fig S7.**
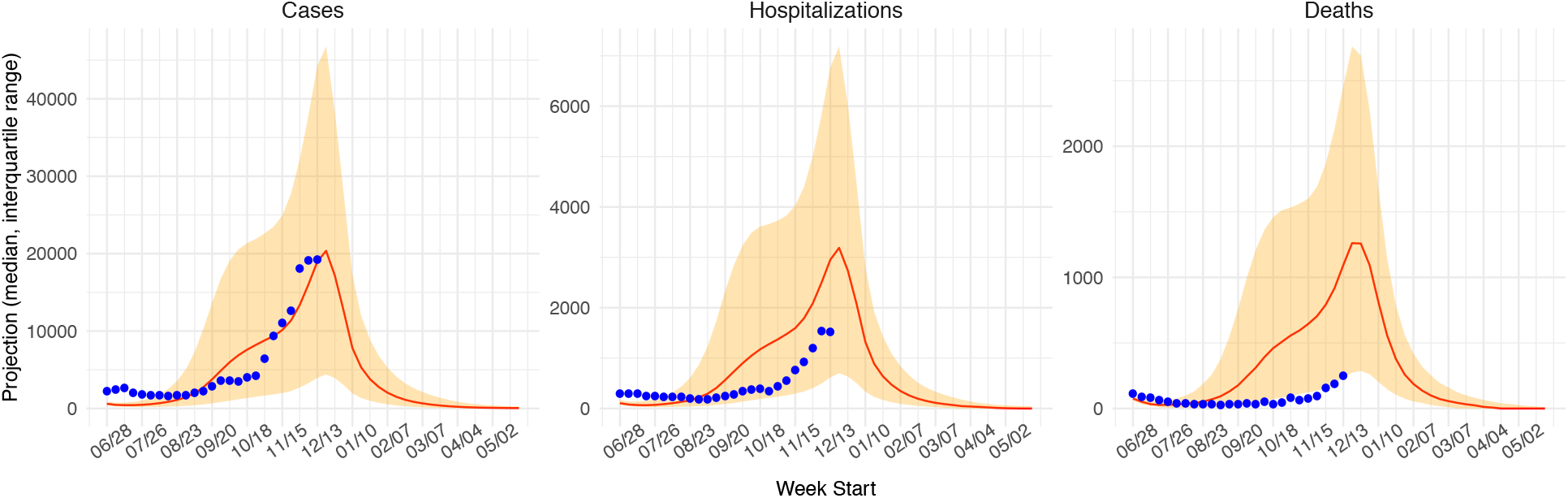
Comparing long term projection with available observations. The projections on COVID-19 cases, hospitalizations and deaths were generated on June 30, 2020, using case and mortality data up to June 26, 2020. These projections were generated at the time using a simpler model without age grouping but otherwise the same methodology presented in the study. Results were posted online on June 30, 2020 (https://github.com/wan-yang/re-opening_analysis/tree/master/test3_occupancy). Various policy scenarios were tested and here we used the one labeled “sce2fix_asIs” for the “intervention” identifier, which has been the closest to implemented interventions thus far (main settings were capping capacity for all industries including schools at 50%; no shutdown). For comparison with observations thus far, we used data published by the New York City Department of Health and Mental Hygiene, as of Dec 27, 2020 (https://raw.githubusercontent.com/nychealth/coronavirus-data/master/trends/data-by-day.csv). Red line and shaded area show our projections (median and interquartile range) and blue dots show corresponding observations from the New York City Department of Health and Mental Hygiene. Note that estimated infection-detection rates using PCR tests during June – Dec 2020 have been similar to those in June. However, hospitalization rates and estimated infection-fatality-risk in months after June 2020 have been lower than that during the spring wave; these lower hospitalization rates and mortality risks contribute to the lower observed numbers than our projections.

